# CytoLens: a modular deep learning system for quantitative cervical cytology

**DOI:** 10.64898/2026.10.13.26365471

**Authors:** Korrine Lee, Rosa Jiwon Yoon, Robyn Sungmi Lee, Dongyong Lee

## Abstract

Cervical cytology screening relies on the visual assessment of nuclear and cellular morphology across thousands of cells per slide, a labor-intensive task that could benefit from quantitative computational support. Here we present CytoLens, a modular deep learning system that detects and counts nuclei, delineates nuclei and whole cells, measures the nuclear-to-cell area ratio (N:C), classifies cells from multifocal images and scores candidate abnormality. We trained and tested each module on a public dataset matched to its task, without pooling datasets. CytoLens detected nuclei with an F1 score of 0.931 (95% confidence interval (CI), 0.914–0.946) and counted 75 of 100 fields to within one nucleus. Point-prompted segmentation achieved mean Dice coefficients of 0.901 for nuclei and 0.847 for whole cells and, without retraining, transferred to a second dataset in which measured N:C increased with lesion grade in parallel with expert outlines. Multifocal hierarchical classification reached a mean accuracy of 0.947 across four Bethesda-aligned categories, and abnormality scores ranked annotated abnormal candidates with an area under the receiver operating characteristic curve (AUROC) of 0.865. These results support the evaluation of CytoLens on patient-linked clinical specimens.

---

Cervical cancer screening by cytology depends on the recognition of a small number of abnormal cells among many normal ones. Cytologists assess nuclear size, the proportion of the cell occupied by the nucleus, chromatin pattern and cell borders, and summarize these observations in the reporting categories of the Bethesda System^1^. The task is labor-intensive, and artificial intelligence (AI) has long been explored as a way to support it; AI-assisted screening systems have now been validated in clinical workflows^2^.

In histopathology, deep learning has reached clinical-grade performance for cancer detection on whole-slide images^3^ and pathologist-level Gleason grading in multinational validation^4^, and foundation models trained on large slide collections now transfer across many diagnostic tasks^5–7^. Most of these systems produce slide-level or tile-level predictions. Cytology interpretation, by contrast, rests on measurable properties of individual cells: how many nuclei a field contains, how large a nucleus is relative to its cell and which morphological category a cell belongs to. An assistant that reports these quantities could be checked directly against what a reader sees, and each quantity could be validated against a reference standard suited to it.

Public cervical cytology datasets make such validation possible because their annotations are complementary. Cx22 provides exhaustive nuclear annotations in fields of overlapping cells^8^; SIPaKMeD^9^ and Herlev^10^ provide nuclear and whole-cell outlines across normal and abnormal categories; CCCID images each cell at 11 focal planes within a two-level morphology hierarchy^11^; and RIVA provides independently annotated candidate cells in conventional Pap smears^12^.

Here we present CytoLens, a modular deep learning system for quantitative cervical cytology (Fig. 1). We evaluated each of its five modules on the dataset that supports its task, at the unit of the reference standard and on group-disjoint test partitions. CytoLens detected and counted nuclei accurately in crowded fields, delineated nuclei and whole cells from a single point prompt, transferred to an unseen dataset without retraining while preserving the relationship between the nuclear-to-cell area ratio (N:C) and lesion grade, and classified multifocal cell stacks with coherent broad-category and fine-category outputs.

**Fig. 1.**
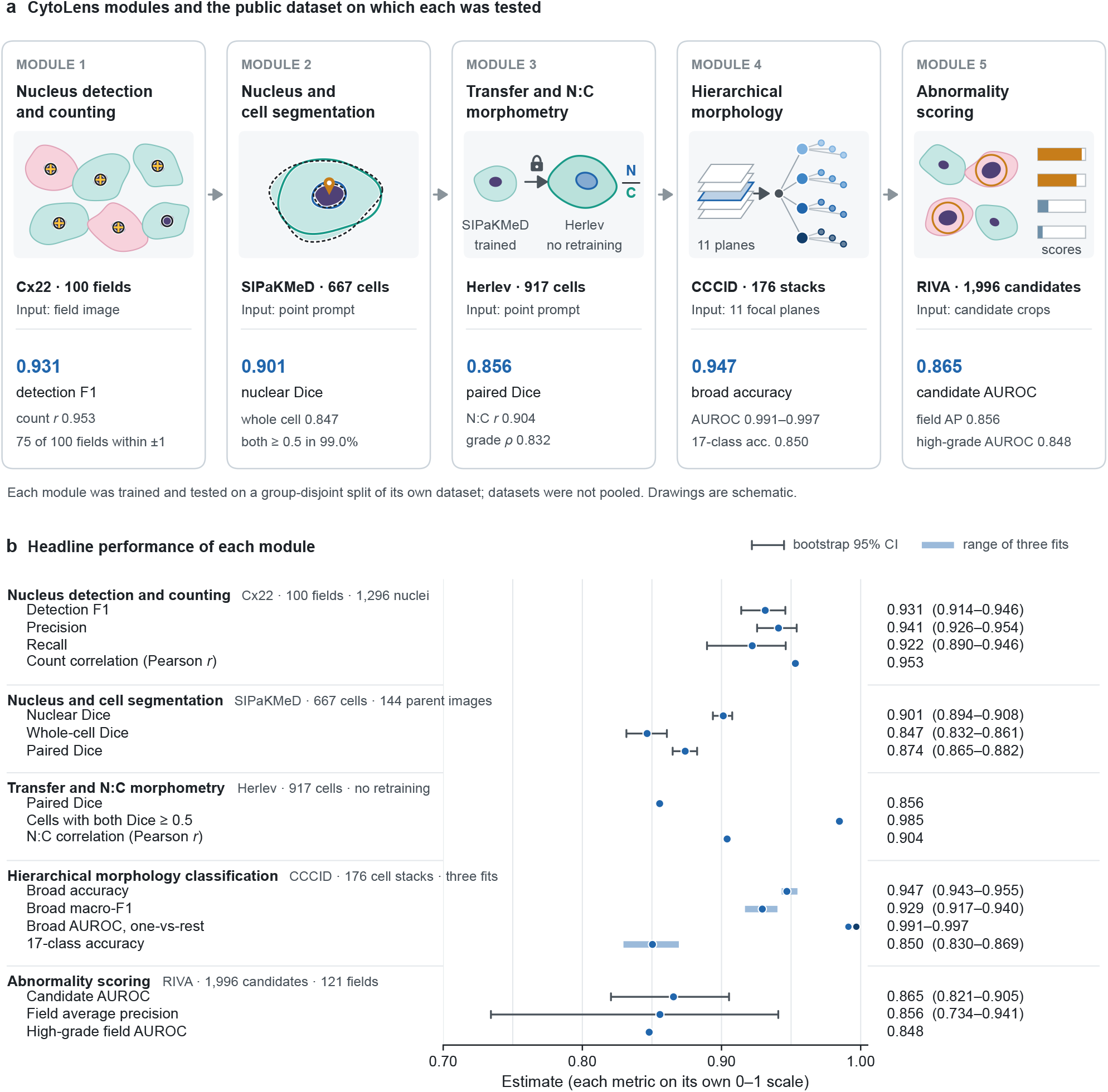
CytoLens modules and headline performance. **a**, The five CytoLens modules in workflow order. Each card names the public dataset on which the module was trained and tested, the input it receives and its headline result. The pin marks the point prompt that indicates the target cell for segmentation. Drawings are schematic and contain no microscopy data. Modules were evaluated separately on group-disjoint test partitions of their own datasets, which were not pooled. **b**, Headline metrics of each module on a common 0–1 axis; each metric keeps its own definition (Tables 2 and 3). Points show estimates, thin lines with caps show bootstrap 95% confidence intervals (CIs; resampling source images for Cx22, parent images for SIPaKMeD and inferred source groups for RIVA) and shaded bars show the range across three independently trained CCCID models, with the mean as the point. The four CCCID AUROC points are one-versus-rest values for NILM, LSIL, HSIL and SCC/other in the primary model (0.991–0.997). Herlev values are descriptive. Field average precision depends on the annotated prevalence (0.545). AUROC, area under the receiver operating characteristic curve; N:C, nuclear-to-cell area ratio.

## Results

We evaluated CytoLens on five public datasets, each matched to one module (Fig. 1a and Table 1). Each module was trained and tested on a group-disjoint split of its own dataset, and datasets were not pooled (Methods). Figure 1b summarizes the headline performance of all five modules.

**Table 1.**
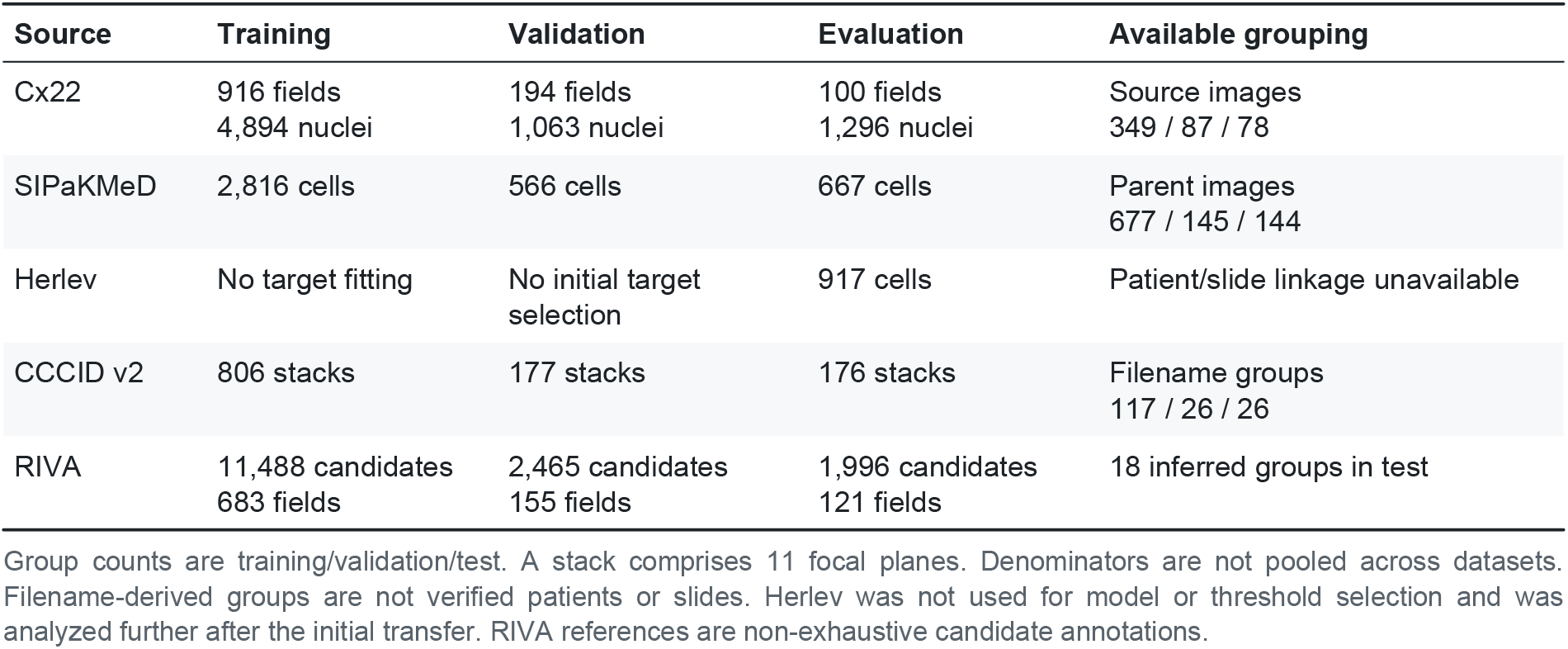
Datasets, partitions and evaluation units.

### CytoLens detects and counts nuclei in crowded fields

The 100 Cx22 test fields contained 1,296 reference nuclei. CytoLens produced 1,270 detections, of which 1,195 matched a reference nucleus, corresponding to a precision of 0.941 (95% confidence interval (CI), 0.926–0.954), a recall of 0.922 (95% CI, 0.890–0.946) and an F1 score of 0.931 (95% CI, 0.914–0.946; Fig. 2a,b and Table 2). Model and reference counts per field were closely correlated (Pearson *r* = 0.953), with a mean absolute error (MAE) of 1.06 nuclei per field (95% CI, 0.78–1.49; Fig. 2c). The model count matched the reference exactly in 42 fields, was within one nucleus in 75 fields and was within two nuclei in 93 fields (Fig. 2d). The mean difference between model and reference counts was −0.26 nuclei per field, with limits of agreement of −4.46 to 3.94 (Extended Data Fig. 1a).

**Fig. 2.**
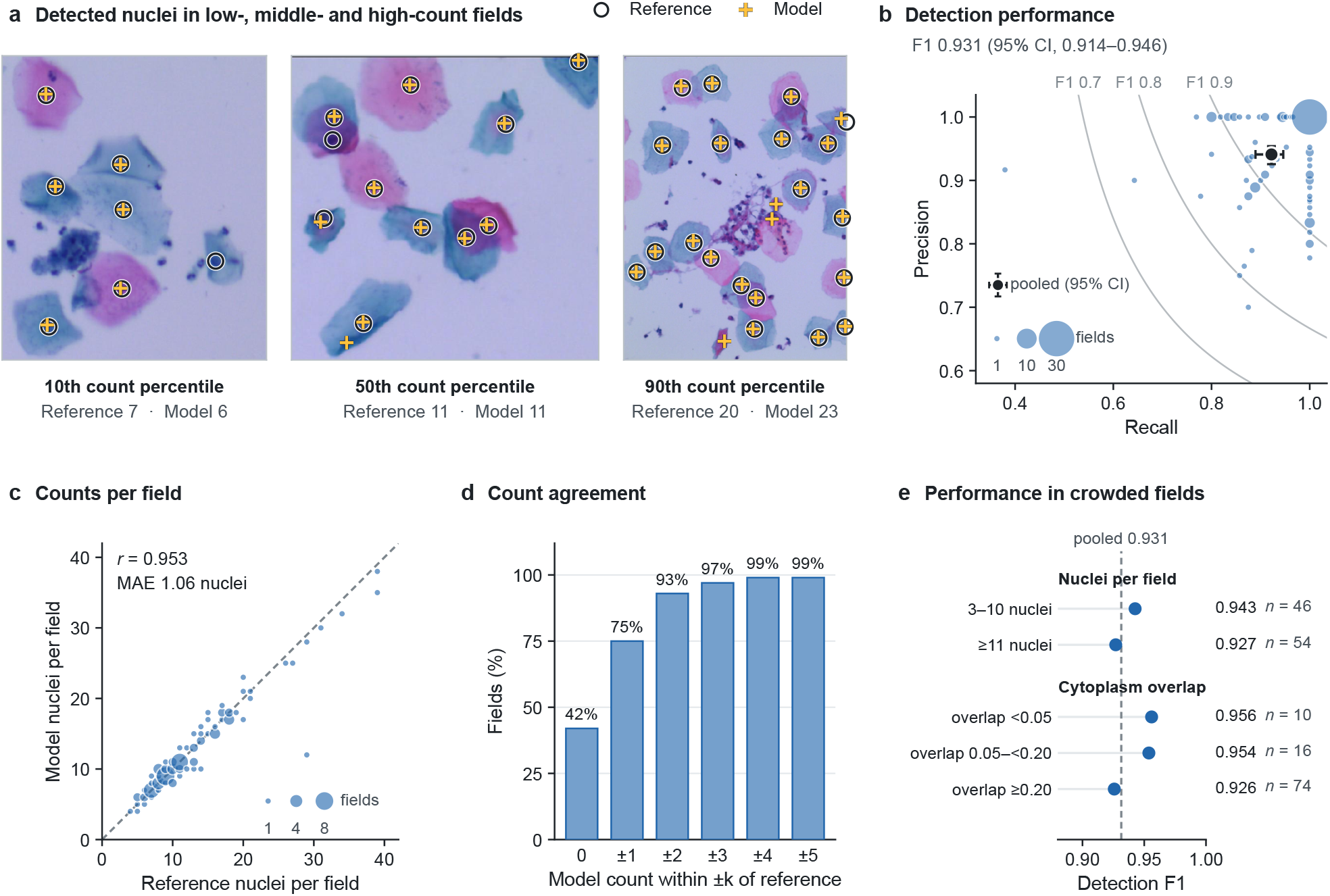
CytoLens detects and counts nuclei accurately. **a**, Three Cx22 test fields at the 10th, 50th and 90th percentiles of the reference nucleus count, selected without reference to model output. Open circles mark reference nuclei and crosses mark CytoLens detections; captions give the reference and model counts. Image content is shown at native aspect ratio (445 × 512, 512 × 507 and 375 × 512 pixels); scale bars are omitted because the release does not record the magnification of each image. **b**, Pooled precision (0.941) and recall (0.922) with bootstrap 95% CIs (black; *n* = 100 fields from 78 source images), giving an F1 score of 0.931 (95% CI, 0.914–0.946), together with per-field values (blue; marker area proportional to the number of fields at each position). Gray curves are lines of constant F1. **c**, Model versus reference nuclei per field (Pearson *r* = 0.953; MAE, 1.06 nuclei; 95% CI, 0.78–1.49); the dashed line shows identity and marker area is proportional to the number of fields. **d**, Percentage of fields in which the model count lies within *k* nuclei of the reference count: 42% exact, 75% within ±1 and 93% within ±2. **e**, Pooled F1 score within strata of nucleus count and cytoplasm-overlap ratio (*n*, number of fields); the dashed line marks the pooled value. In the most overlapped fields (ratio ≥0.20; *n* = 74), F1 was 0.926. MAE, mean absolute error.

**Table 2.** Performance of the detection, segmentation, morphometry and classification modules.

| Endpoint | Evaluation unit | Estimate | Uncertainty or context |
| --- | --- | --- | --- |
| Nucleus detection F1 score | Cx22: 100 fields | 0.931 | 95% CI, 0.914–0.946 |
| Precision / recall | Cx22: 100 fields | 0.941 / 0.922 | 95% CI, 0.926–0.954 / 0.890–0.946 |
| Nucleus count MAE | Cx22: 100 fields | 1.06 per field | 95% CI, 0.78–1.49 |
| Count correlation; fields within $\pm 1$ nucleus | Cx22: 100 fields | $r = 0.953$; 75% | 93% within $\pm 2$ |
| Nuclear / whole-cell Dice | SIPaKMeD: 667 cells | 0.901 / 0.847 | 95% CI, 0.894–0.908 / 0.832–0.861 |
| Paired Dice | SIPaKMeD: 667 cells | 0.874 | 95% CI, 0.865–0.882 |
| Cells with both Dice $\geq 0.5$ / $\geq 0.7$ | SIPaKMeD: 667 cells | 99.0% / 88.9% | 95% CI, 97.7%–99.8% ($\geq 0.5$) |
| Paired Dice without retraining | Herlev: 917 cells | 0.856 | Nuclear 0.878; whole cell 0.834 |
| Cells with both Dice $\geq 0.5$ / $\geq 0.7$ | Herlev: 917 cells | 98.5% / 90.8% | Descriptive |
| N:C correlation / MAE | Herlev: 917 cells | 0.904 / 0.097 | Mean difference $-0.085$; limits $-0.284$ to $0.114$ |
| N:C rank correlation with grade | Herlev: 819 squamous cells | 0.832 | Reference outlines 0.837; exploratory |
| Broad accuracy | CCCID: 176 stacks | 0.947 | Three-fit range 0.943–0.955 |
| Broad macro-averaged F1 score | CCCID: 176 stacks | 0.929 | Three-fit range 0.917–0.940 |
| Broad one-versus-rest AUROC | CCCID: 176 stacks | 0.991–0.997 | Fit 641; four groups |
| 17-class accuracy / macro-averaged F1 score | CCCID: 176 stacks | 0.850 / 0.820 | Ranges 0.830–0.869 / 0.783–0.845 |
| Fine prediction within the predicted broad group | CCCID: 176 stacks | 176 / 176 | Fit 641 |

Confidence intervals (CIs) resample source images (Cx22) or parent images (SIPaKMeD); ranges across the three CCCID models are not CIs; Herlev values are descriptive because the release has no grouping variable. The segmentation model receives a point prompt at the nuclear centroid of the target cell, so Dice values describe delineation of an indicated cell and are not directly comparable with results obtained without a prompt. Paired Dice is the mean of nuclear and whole-cell Dice coefficients. AUROC, area under the receiver operating characteristic curve; MAE, mean absolute error; N:C, nuclear area divided by whole-cell area (nucleus included).

Performance was maintained in crowded and overlapping fields. The F1 score was 0.943 in fields with 3–10 nuclei and 0.927 in fields with 11 or more, and 0.956, 0.954 and 0.926 in fields with low, intermediate and high cytoplasm overlap, respectively (Fig. 2e). The high-overlap stratum comprised 74 of the 100 test fields, and count errors were only weakly associated with overlap (Spearman *ρ* = 0.178; Extended Data Fig. 1b).

### A single point prompt yields accurate nuclear and whole-cell boundaries

Given a point prompt at the nucleus of a target cell, CytoLens delineated the nucleus and the whole cell in 667 SIPaKMeD test cells from 144 parent images with mean Dice coefficients of 0.901 (95% CI, 0.894–0.908) and 0.847 (95% CI, 0.832–0.861), respectively, and a mean paired Dice coefficient of 0.874 (95% CI, 0.865–0.882; Fig. 3a,b). Both compartments reached a Dice coefficient of at least 0.5 in 99.0% of cells (95% CI, 97.7–99.8%) and at least 0.7 in 88.9% (Fig. 3c). Performance was consistent across the five source classes, with mean nuclear Dice coefficients of 0.871–0.928 and success rates of 97.8–100% at the 0.5 threshold (Fig. 3c and Extended Data Fig. 2a,b), and across parent images (Extended Data Fig. 2c,d).

**Fig. 3.**
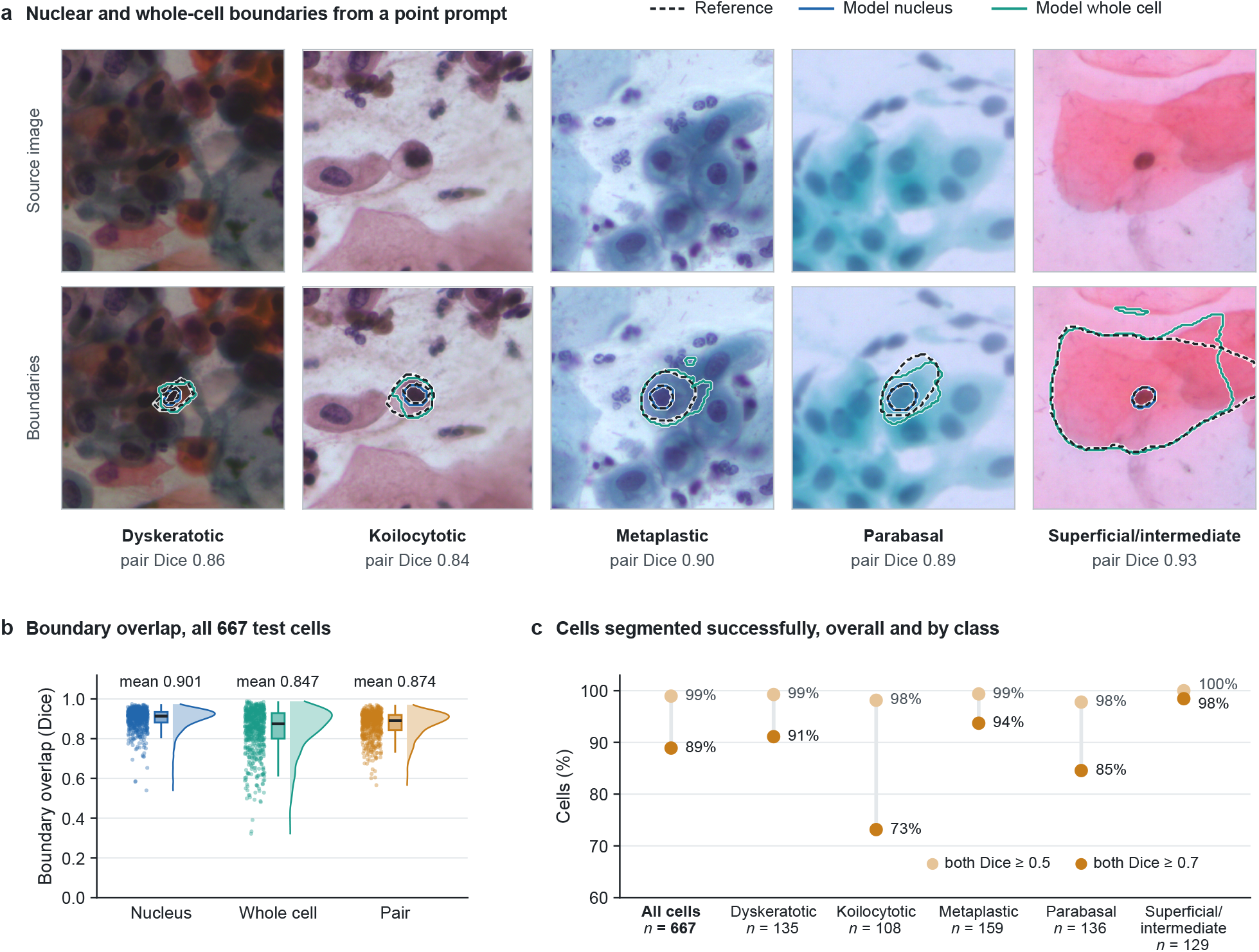
Point-prompted segmentation delineates nuclei and whole cells. **a**, One SIPaKMeD test cell per source class, shown as the original 512-pixel source window (top) and with reference (black dashed), CytoLens nuclear (blue) and whole-cell (teal) contours (bottom). Examples lie near the within-class median paired Dice coefficient, given in each caption. Contours are the evaluated masks mapped to the source window; for the koilocytotic cell, 16 rows of white padding are omitted. The provider does not report pixel size, so scale bars are omitted. **b**, Nuclear (blue), whole-cell (teal) and paired (amber) Dice coefficients for all test cells (*n* = 667 cells from 144 parent images), each shown as individual cells, a density estimate and a box plot. Means (parent-image bootstrap 95% CIs) are 0.901 (0.894–0.908), 0.847 (0.832–0.861) and 0.874 (0.865–0.882); paired Dice is the mean of the two compartment coefficients. **c**, Percentage of cells in which both the nuclear and the whole-cell mask reached a Dice coefficient of at least 0.5 (light) or 0.7 (dark), overall (99.0% and 88.9%) and per class. Boxes indicate quartiles and whiskers extend to the most extreme values within 1.5 × the interquartile range.

### Segmentation transfers without retraining and preserves the N:C gradient

We next applied the SIPaKMeD-trained model, unchanged, to all 917 cells of the Herlev dataset. Mean nuclear, whole-cell and paired Dice coefficients were 0.878, 0.834 and 0.856, respectively, and both compartments reached a Dice coefficient of at least 0.5 in 98.5% of cells and at least 0.7 in 90.8% (Fig. 4a). Relative to an earlier Cx22-trained version of the pipeline, the mean paired Dice coefficient increased from 0.381 to 0.856, with improvement in 99% of cells (Extended Data Fig. 3c,d).

**Fig. 4.**
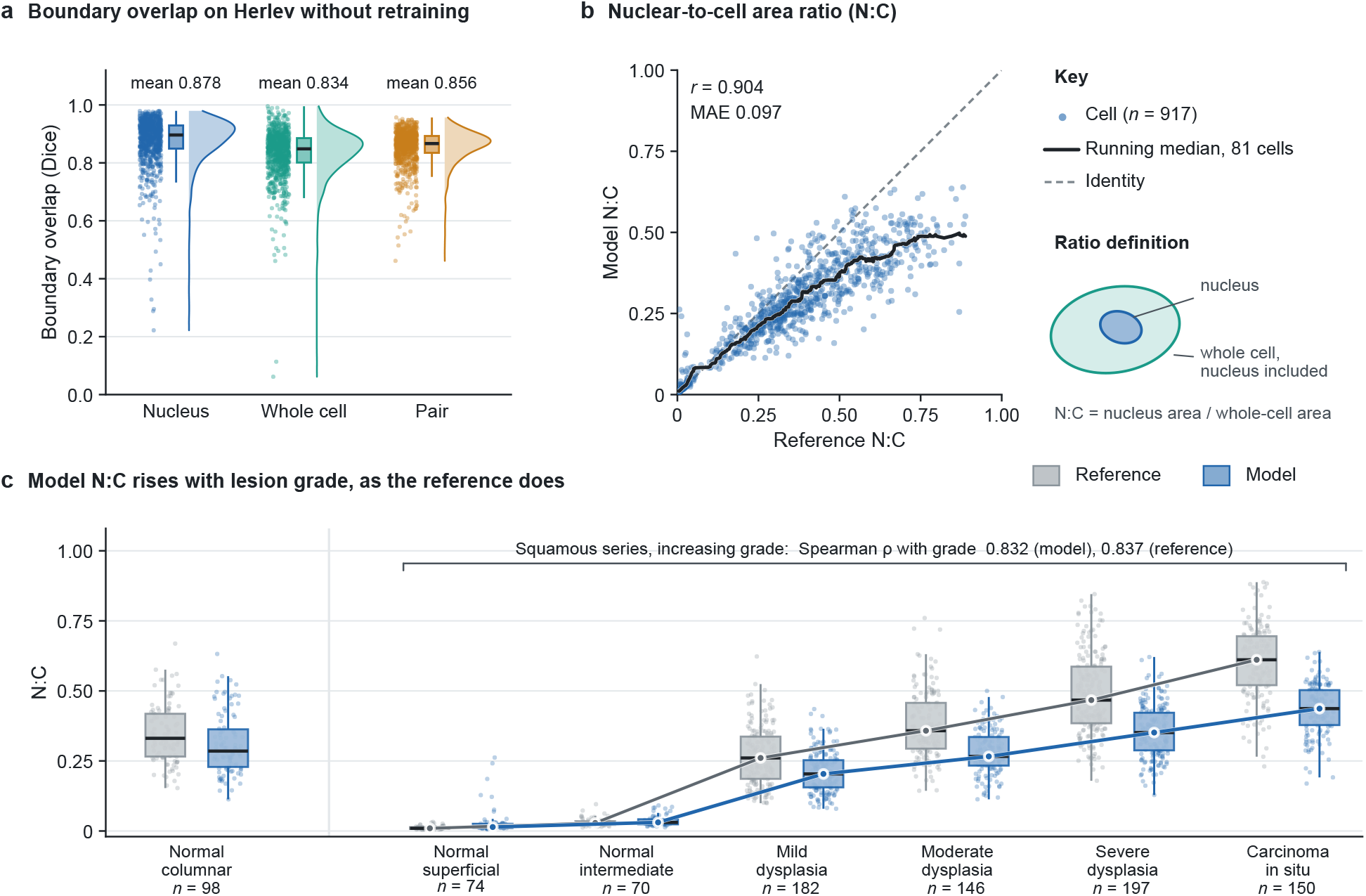
The segmentation model transfers to Herlev without retraining and preserves the N:C gradient. **a**, Nuclear, whole-cell and paired Dice coefficients for all Herlev cells (*n* = 917), obtained with the SIPaKMeD-trained model, input window and mask thresholds unchanged. Means are 0.878, 0.834 and 0.856; both compartments reached a Dice coefficient of at least 0.5 in 98.5% of cells and at least 0.7 in 90.8%. **b**, Model versus reference N:C for every cell, with the identity line (dashed) and an 81-cell running median (black); Pearson *r* = 0.904; MAE, 0.097. Model ratios are lower than the reference by 0.085 on average (Extended Data Fig. 3a). **c**, Reference (gray) and model (blue) N:C by Herlev class. Lines join class medians across the squamous series (normal superficial and intermediate, mild, moderate and severe dysplasia, carcinoma in situ; *n* = 819 cells), in which the Spearman correlation of N:C with grade was 0.832 for the model and 0.837 for the reference (exploratory analysis). Normal columnar cells are shown separately. Boxes indicate quartiles and whiskers extend to the most extreme values within 1.5 × the interquartile range. N:C, nuclear area divided by whole-cell area (nucleus included).

The N:C derived from CytoLens masks correlated with the N:C derived from reference outlines (*r* = 0.904; MAE, 0.097; Fig. 4b), with a mean difference of −0.085 (limits of agreement, −0.284 to 0.114; Extended Data Fig. 3a). Across the squamous series, the median model N:C increased at each step of lesion grade, from 0.014 and 0.031 in normal superficial and intermediate cells to 0.204, 0.266, 0.351 and 0.437 in mild, moderate and severe dysplasia and carcinoma in situ, respectively (Fig. 4c). The reference medians followed the same order (0.010, 0.029, 0.261, 0.358, 0.467 and 0.611), and the rank correlation of N:C with grade was 0.832 for the model and 0.837 for the reference outlines in 819 squamous cells. Used alone to separate the four abnormal classes from the three normal classes, model N:C achieved an area under the receiver operating characteristic curve (AUROC) of 0.803, compared with 0.854 for the reference ratio (Extended Data Fig. 3e).

### Hierarchical classification of multifocal cell stacks is accurate and coherent

The CCCID broad categories are negative for intraepithelial lesion or malignancy (NILM), low-grade squamous intraepithelial lesion (LSIL), high-grade squamous intraepithelial lesion (HSIL) and squamous cell carcinoma or other malignancy (SCC/other). On 176 CCCID test stacks, broad-category accuracy averaged 0.947 (range, 0.943–0.955) and the macro-averaged F1 score averaged 0.929 (range, 0.917–0.940) across three independently trained models (Table 2). In the primary model, 84 of 89 NILM, 23 of 24 LSIL, 21 of 23 HSIL and 38 of 40 SCC/other stacks were assigned to the correct category (Fig. 5b), and the one-versus-rest AUROCs were 0.997, 0.996, 0.991 and 0.997, respectively (Fig. 5c). Accuracy across the 17 fine categories averaged 0.850 (range, 0.830–0.869), with a macro-averaged F1 score of 0.820 (range, 0.783–0.845; Extended Data Fig. 4).

**Fig. 5.**
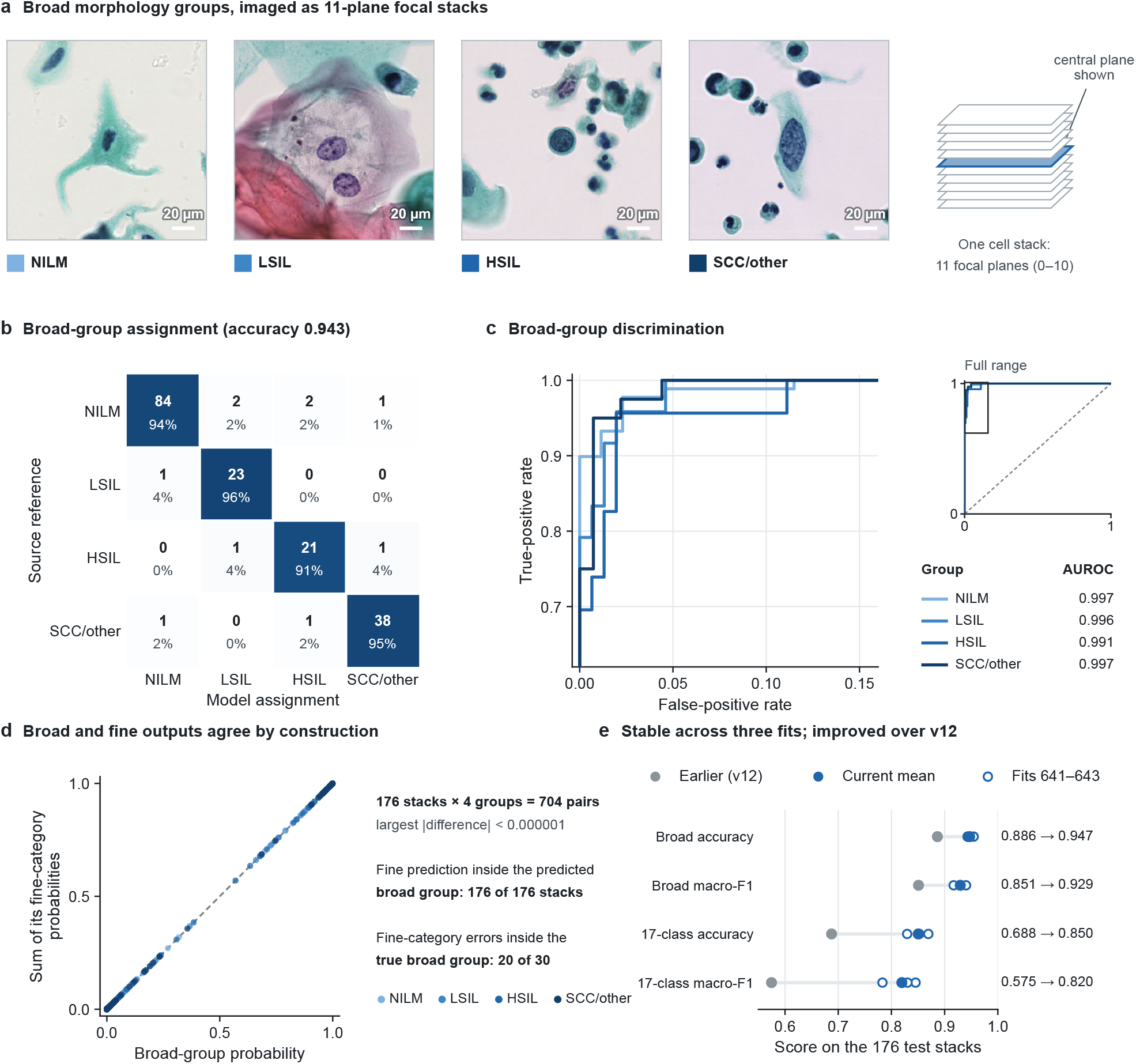
CytoLens classifies multifocal cell stacks with coherent broad and fine outputs. **a**, Central focal plane of one CCCID test stack per broad category (scale bars, 20 µm); each stack comprises 11 focal planes. **b**, Confusion matrix of the primary model on the 176 test stacks from 26 filename groups (counts and row percentages; accuracy, 0.943). **c**, One-versus-rest receiver operating characteristic curves of the primary model at false-positive rates up to 0.16, with the full range inset; AUROCs are 0.997 (NILM), 0.996 (LSIL), 0.991 (HSIL) and 0.997 (SCC/other). **d**, Broad-category probability versus the sum of the probabilities of its fine categories for every test stack and category (*n* = 704), colored by category. All points lie on the identity line (largest absolute difference, <0.000001). The predicted fine category fell within the predicted broad category in 176 of 176 stacks, and 20 of 30 fine-category errors remained within the correct broad category. **e**, Accuracy and macro-averaged F1 score at the broad and 17-class levels for the earlier CytoLens classifier (gray) and the three current models (open circles; filled circles, mean). Current means are 0.947 (range, 0.943–0.955), 0.929 (0.917–0.940), 0.850 (0.830–0.869) and 0.820 (0.783–0.845). The current model was adopted under a prespecified validation protocol (validation gain in 17-class macro-averaged F1, 0.169; 95% CI, 0.054–0.221); test differences are descriptive. AUROC, area under the receiver operating characteristic curve.

The two output levels were coherent. In the primary model, each broad-category probability equaled the sum of its fine-category probabilities to within 0.000001, as guaranteed by the probability composition (Methods); the predicted fine category fell within the predicted broad category in all 176 stacks; and 20 of the 30 fine-category errors remained within the correct broad category (Fig. 5d). The current model outperformed the earlier CytoLens classifier on all four summary measures (Fig. 5e). It had been adopted under a prespecified validation protocol, with a mean gain in the fine-category macro-averaged F1 score of 0.169 (95% CI, 0.054–0.221) on the validation partition; test-set differences are therefore descriptive.

### Abnormality scores rank abnormal candidates and fields

On 1,996 RIVA test candidates, the candidate abnormality score achieved an AUROC of 0.865 (95% CI, 0.821–0.905) and an average precision of 0.751 at a candidate prevalence of 0.320 (Fig. 6a and Table 3). Field scores were higher in the 66 fields containing an annotated abnormal candidate than in the 55 fields without one (Fig. 6b), with a field-level AUROC of 0.803 and an average precision of 0.856 (95% CI, 0.734–0.941) at a prevalence of 0.545 (Fig. 6c,d). For fields containing an annotated high-grade candidate (atypical squamous cells, cannot exclude HSIL (ASC-H); HSIL; or squamous cell carcinoma (SCC)), the high-grade score achieved an AUROC of 0.848 and an average precision of 0.713 at a prevalence of 0.322. At operating points fixed on validation data, the abnormality score had a sensitivity of 0.833 and a specificity of 0.527, and the high-grade score had a sensitivity of 0.692 and a specificity of 0.805 (Table 3 and Extended Data Fig. 5b,c). Assignment of individual candidates to eight categories was less accurate (macro-averaged F1 score, 0.265; Extended Data Fig. 5d), which supports the use of the abnormality score as a ranking signal.

**Fig. 6.**
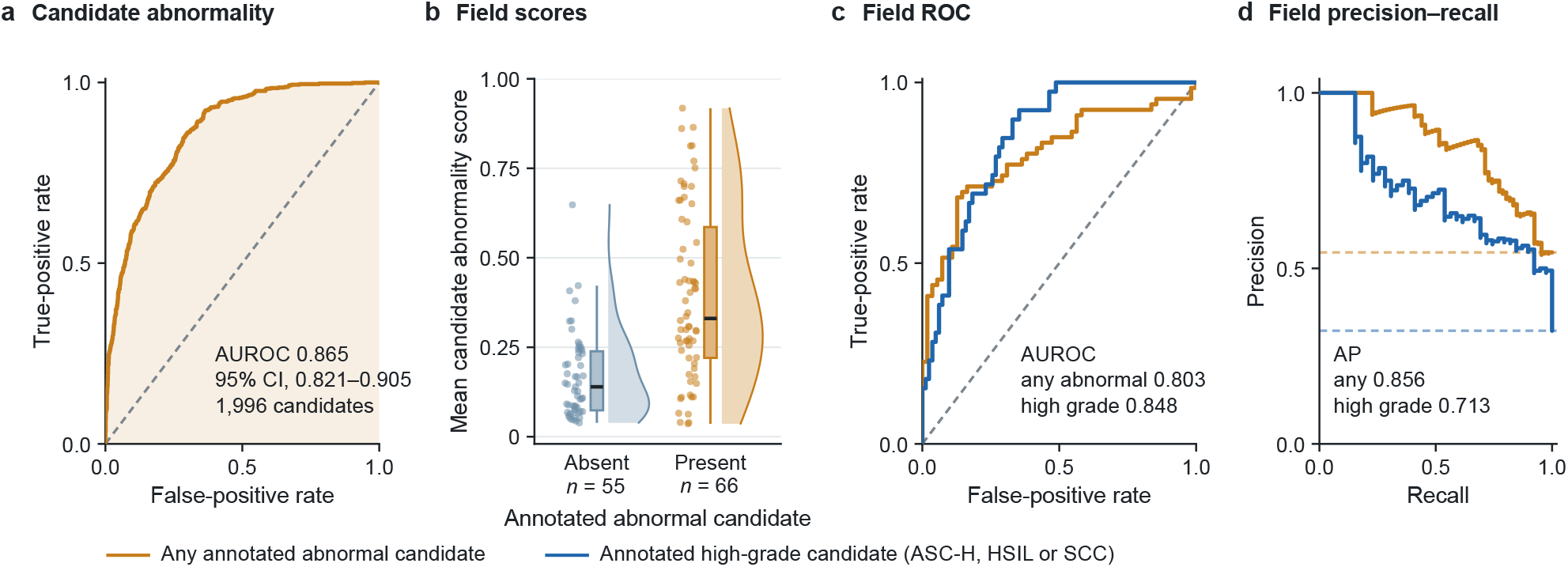
CytoLens abnormality scores rank abnormal candidates and fields. **a**, Receiver operating characteristic (ROC) curve of the candidate abnormality score (summed ASC-US, LSIL, ASC-H, HSIL and SCC probabilities) for RIVA test candidates (*n* = 1,996 from 18 inferred source groups); AUROC, 0.865 (95% CI, 0.821–0.905; inferred-group bootstrap). **b**, Field score (mean candidate abnormality probability) in fields without (*n* = 55) and with (*n* = 66) an annotated abnormal candidate. Boxes indicate quartiles and whiskers extend to the most extreme values within 1.5 × the interquartile range. **c**, Field ROC curves for any annotated abnormal candidate (amber; AUROC, 0.803) and for an annotated high-grade candidate (ASC-H, HSIL or SCC; blue; AUROC, 0.848; *n* = 39 positive fields). **d**, Field precision–recall curves for the same endpoints; average precision 0.856 (95% CI, 0.734–0.941) and 0.713. Dashed lines mark the annotated prevalences (0.545 and 0.322), on which precision depends. Scores, aggregation and thresholds were fixed on validation data. RIVA annotations are non-exhaustive, so field labels are not slide-level or patient-level diagnoses. AUROC, area under the ROC curve.

**Table 3.** Abnormality scoring on RIVA.

| Endpoint | Value | Evaluation context |
| --- | --- | --- |
| Candidate abnormality AUROC | 0.865 (0.821–0.905) | 1,996 candidates; inferred-group 95% CI |
| Candidate abnormality average precision | 0.751 | Candidate prevalence 0.320 |
| Field abnormality average precision | 0.856 (0.734–0.941) | 121 fields; prevalence 0.545; inferred-group 95% CI |
| Field abnormality AUROC | 0.803 | 66 positive / 55 negative annotated fields |
| High-grade field AUROC / average precision | 0.848 / 0.713 | 39 positive / 82 negative fields; prevalence 0.322 |
| Field abnormality sensitivity / specificity | 0.833 / 0.527 | Threshold fixed on validation (0.149) |
| Field abnormality TP / FP / FN / TN | 55 / 26 / 11 / 29 | Same fixed threshold |
| High-grade sensitivity / specificity | 0.692 / 0.805 | Threshold fixed on validation (0.112) |
| High-grade TP / FP / FN / TN | 27 / 16 / 12 / 66 | Same fixed threshold |
| Eight-category candidate macro-averaged F1 score | 0.265 | Fine candidate assignment |

Abnormality comprises ASC-US, LSIL, ASC-H, HSIL and SCC; high grade comprises ASC-H, HSIL and SCC. Field scores average candidate probabilities; aggregation and thresholds were fixed on validation data before testing. RIVA annotations are non-exhaustive and annotation-level, so field labels are not slide or patient diagnoses, and average precision depends on the annotated prevalence. AUROC, area under the receiver operating characteristic curve; CI, confidence interval; TP/FP/FN/TN, true-positive/false-positive/false-negative/true-negative field counts.

## Discussion

CytoLens provides accurate, quantitative readouts of cervical cytology at the level at which cytologists reason: nuclei, cell boundaries, nuclear-to-cell proportion and cell category. Across five public datasets, it detected nuclei with an F1 score of 0.931, delineated nuclei and whole cells from a single point prompt, transferred to a new dataset without retraining and classified multifocal cell stacks into Bethesda-aligned categories with a mean accuracy of 0.947.

The N:C measurements illustrate why such readouts matter. Nuclear enlargement relative to the cell is a central criterion for grading squamous lesions^1^, and CytoLens reproduced the stepwise increase in N:C from normal squamous cells to carcinoma in situ on a dataset that it had not seen during training, with a rank correlation with grade almost identical to that of expert outlines. The small systematic offset relative to the reference (−0.085) left this ordering intact and could be removed by site-specific calibration.

The hierarchical classifier addresses a practical requirement of reader-facing tools, namely that outputs at different levels of detail should not contradict each other. Because fine-category probabilities are composed within their broad parent, the two levels agree by construction. In the primary model, every fine prediction fell within the predicted broad category, and most fine-category errors were confusions between related subtypes rather than between broad categories. A display can therefore present a broad call together with its finer differential without inconsistency.

Several observations indicate that this performance is robust rather than confined to favorable subsets. Detection remained accurate in the most overlapped fields, which made up three-quarters of the test set; segmentation retained most of its accuracy on an unseen dataset; and broad-category accuracy varied by about one percentage point across independently trained models. Iterative development also produced measurable gains under prespecified criteria: the current classifier improved the fine-category macro-averaged F1 score from 0.575 to 0.820, and the current segmentation pipeline raised the paired Dice coefficient on Herlev from 0.381 to 0.856.

These component-level results complement workflow-level validation of AI-assisted screening^2^ and the slide-level advances of computational pathology^3–7^. They establish which quantities an assistant measures accurately and on which data. Because CytoLens segmentation receives a point prompt, its Dice coefficients are not directly comparable with published SIPaKMeD or Herlev results obtained without one; a matched comparison would require a common frozen split, identical inputs and equal model-selection budgets. The modules use standard architectures and are fully specified, so they can serve as a baseline for such comparisons.

Our study has several limitations. First, it is a retrospective analysis of public data. Cx22 and RIVA were examined repeatedly during development, and Herlev was analyzed further after the initial transfer, so the reported intervals may understate uncertainty. Second, patient identifiers and biopsy outcomes were unavailable; filename-derived groups are not verified patients, and the small number of groups in some test partitions (18 for RIVA and 26 for CCCID) widens the clustered intervals. Third, the modules were evaluated separately. Segmentation used the reference nuclear centroid as its prompt, so the effect of missed or spurious detections on downstream measurements remains to be measured. Fourth, model N:C was on average 0.085 lower than the reference on Herlev, fine-category performance was lower than broad-category performance, and the RIVA annotations are non-exhaustive, with an enriched field prevalence and a specificity of 0.527 at the fixed abnormality threshold. Finally, demographic, scanner-stratified and reader-level analyses were not possible with the available metadata, and we did not perform a matched comparison with current foundation models.

In summary, CytoLens provides accurate and interpretable measurements of nuclei, cell boundaries, nuclear-to-cell proportion and cell category across five public cervical cytology datasets. Evaluation of the integrated pipeline on newly acquired, patient-linked specimens, together with site calibration of morphometric outputs and a reader study, is warranted before clinical use.

## Methods

### Study design and ethics

This retrospective computational study used only openly available, de-identified cervical cytology images and annotations, obtained from Cx22 (https://github.com/LGQ330/Cx22), SIPaKMeD (https://www.cs.uoi.gr/∼marina/sipakmed.html), Herlev (https://mde-lab.aegean.gr/index.php/downloads/), CCCID version 2 (https://doi.org/10.5281/zenodo.20807462), RIVA (https://zenodo.org/records/17288879) and APACC (https://osf.io/fp2xe). No new participants were recruited, no identifiable patient records were accessed and no intervention was performed. The original collections were obtained under the ethical approvals and consent procedures documented by their providers^8–12,14^. Because only openly available, de-identified data were analyzed, no additional ethics approval was sought for this secondary analysis. The study is reported in accordance with the 2024 update of the Checklist for Artificial Intelligence in Medical Imaging^13^ and was not prospectively registered.

The model-development experiments were executed before preparation of this manuscript. Their configuration files, split manifests, selection records, prediction tables and checkpoint hashes were used to reconstruct every reported value. Checkpoints and thresholds were selected on the recorded validation partitions; no model was refitted and no threshold was reoptimized for this report. The descriptive analyses described under Statistical analysis were computed from the same frozen predictions. Data were drawn from the available public releases rather than from a sample-size calculation.

### Datasets and partitions

The Cx22 analysis used 916 training fields, 194 validation fields and 100 test fields from 349, 87 and 78 source-image groups, respectively, containing 4,894, 1,063 and 1,296 reference nuclei (Table 1). Development images sharing source identifiers with the test partition were excluded before fitting. The test partition comprised the designated multi-cell test subset, in which the source masks provide exhaustive nuclear references.

SIPaKMeD contained 4,049 cells linked to 966 parent images. The grouped split comprised 2,816 training cells from 677 parents, 566 validation cells from 145 parents and 667 test cells from 144 parents. A 512-pixel square window centered on the nuclear centroid was resized to 128 × 128 pixels; image and polygon coordinates were transformed together, and locations outside the original image were filled with white. Whole-cell reference masks included the nucleus.

Herlev contributed all 917 image–mask pairs across its seven legacy categories^10^. The SIPaKMeD-trained segmentation model, its 512-pixel input window and its mask thresholds were applied without any Herlev-based training or selection. The labels were treated as source annotations rather than as patient-level biopsy end points.

The CCCID version 2 main release contained 1,159 complete cell stacks, each with 11 focal planes, in 169 conservative filename-derived groups^11^. The separately packaged AI subset was not counted as additional data. The frozen split contained 806 training, 177 validation and 176 test stacks from 117, 26 and 26 groups, respectively. Each focal-plane image was resized to 224 × 224 pixels. The four broad labels were NILM, LSIL, HSIL and SCC/other, the last including malignant entities other than squamous cell carcinoma, and 17 source-defined fine categories were retained within this hierarchy. Test broad-category supports were 89, 24, 23 and 40 stacks.

The RIVA analysis used the annotated region and candidate release^12^. The frozen candidate prediction export contained 11,488 training, 2,465 validation and 1,996 test candidates in 683, 155 and 121 fields, respectively; 18 inferred groups were represented in the test set. Because annotations may be non-exhaustive, the absence of an annotated abnormal candidate was not equated with a negative slide, and no unannotated location was assigned a new label.

### Evaluation units and partitioning

Each end point was computed at the unit provided by its reference standard: fields for Cx22; cells for SIPaKMeD and Herlev; 11-plane stacks for CCCID; and candidates or fields for RIVA. Datasets were never pooled into a single sample size, accuracy or CI, and dataset-specific labels were kept as released. Partitions were disjoint by the strongest grouping variable that each release supplies, and the same variable was used for clustered uncertainty: source images for Cx22, parent images for SIPaKMeD and filename-derived groups for CCCID and RIVA. Filename-derived groups are not verified patients, and Herlev, which provides no grouping variable, is reported descriptively. The input of each module is reported with its result, and every operating point was fixed on validation data before testing.

### Nucleus detection and counting

A point-heatmap U-Net^15^ received 224 × 224-pixel Cx22 images. The implementation used 24 base channels and a center-focal loss against nuclear-center heatmaps. Training used AdamW, a batch size of 8, a learning rate of 0.0003, a weight decay of 0.0001, gradient-norm clipping at 5 and at most 10 epochs. Dihedral and modest brightness, contrast and saturation augmentations were applied to training images only, and sampling accounted for cell count and overlap.

The validation-selected peak threshold was 0.4 and the non-maximum-suppression radius was 5 model-input pixels. Reference and predicted centers were matched one-to-one within a 7-pixel radius in the model-input coordinate system, accepting eligible pairs greedily in ascending order of distance. Precision, recall and F1 score pooled matched counts across the evaluated fields. The field count was the number of retained peaks, and count agreement was summarized by the MAE, the Pearson correlation coefficient, Bland–Altman limits of agreement and the percentage of fields counted within *k* nuclei of the reference.

### Point-prompted segmentation

A point-prompted paired U-Net received the RGB crop together with a Gaussian point-prompt channel (s.d., 4 pixels) marking the nucleus of the target cell and produced separate nuclear and whole-cell masks. The model used 24 base channels and was initialized from an earlier Cx22-trained center-prompted paired model. Adaptation to SIPaKMeD used AdamW with a learning rate of 0.0002, a weight decay of 0.0001, a batch size of 32 and at most 14 epochs. The loss combined class-balanced binary cross-entropy and soft Dice loss for each compartment, a nuclear-containment penalty and a whole-cell boundary term, weighted 1.25 for the nucleus, 1.0 for the whole cell, 0.25 for containment and 0.15 for the boundary.

Checkpoint selection prioritized the validation percentage of cells with both compartments at a Dice coefficient of at least 0.5, followed by the mean paired Dice coefficient; both mask thresholds were set to 0.75 on validation data. Nuclear and whole-cell Dice coefficients were reported separately, and the paired Dice coefficient was their arithmetic mean. In this evaluation, the point prompt was placed at the reference nuclear centroid, so the results describe delineation of an indicated cell; in use, the prompt can come from the detection module or from a reader selecting a cell.

### N:C morphometry and transfer to Herlev

N:C was defined as the nuclear mask area divided by the whole-cell mask area. Because the whole-cell mask includes the nucleus, this ratio differs from a nucleus-to-cytoplasm ratio. Reference and model ratios were computed on the same cells. To test transfer, the SIPaKMeD-trained model, input window and thresholds were applied unchanged to all 917 Herlev cells, with Herlev reference nuclear centroids as point prompts.

### Hierarchical multifocal classifier

The CCCID model used an ImageNet-initialized ResNet-18 encoder^16^ with a four-class broad-category head, a 17-class fine-category head, an ordinal head and a 128-dimensional projection head. During training, two focal planes from the same cell stack formed a positive pair, and training-only cross-focus pretraining used a symmetric contrastive loss^17^ for six epochs, with a learning rate of 0.00005 and a temperature of 0.15. Supervised training combined a broad-category balanced softmax loss^18^, a within-parent conditional fine-category loss, an ordinal binary cross-entropy loss and the cross-focus contrastive loss, weighted 0.7, 0.35 and 0.08 relative to the broad-category term. Training used a batch size of 32, a learning rate of 0.00008, a weight decay of 0.0001, gradient clipping at 5 and at most 16 epochs, with sampling that compensated for broad-category and filename-group representation.

To enforce ordered cumulative outputs, the first ordinal logit was unconstrained and each subsequent logit was obtained by subtracting a softplus-transformed network output, which yields non-increasing cumulative probabilities; this follows the principle of rank-consistent ordinal regression^19^ without reimplementing it. Broad-category probabilities were a convex mixture of nominal softmax probabilities and ordinal probabilities. Within each broad category, a softmax over its permitted fine categories was multiplied by the broad-category probability, so that fine-category probabilities sum to their parent probability by construction.

At inference, the logits of each head were averaged over all 11 planes before probability composition. The checkpoint was selected by the validation fine-category macro-averaged average precision, with the fine-category macro-averaged F1 score as a tiebreaker, and the ordinal mixture coefficient was selected from 0, 0.25, 0.5, 0.75 and 1 on validation data (0.25 for the primary model, seed 641). Three models were trained with seeds 641–643 on the same frozen split; confusion matrices and receiver operating characteristic (ROC) curves are shown for the primary model, and the three-model mean is not an ensemble. The model replaced an earlier CytoLens classifier (a ResNet-18 multitask model without hierarchical composition, ordinal head or cross-focus pretraining) under a prespecified validation protocol, which required a gain in the fine-category macro-averaged F1 score with a 95% CI excluding zero and noninferiority at the broad-category level; the test set was not used for adoption.

### Abnormality scoring

The RIVA classifier combined three ResNet-18-based models, a predecessor hierarchical model and two factorized variants, with validation-selected probability weights of 0.50, 0.25 and 0.25 and an archived temperature of 0.8483953846. The candidate abnormality score was the sum of the probabilities for atypical squamous cells of undetermined significance (ASC-US), LSIL, ASC-H, HSIL and SCC; NILM, inflammatory and endocervical labels formed the complementary group. A separate high-grade score summed the ASC-H, HSIL and SCC probabilities. The field score was the arithmetic mean of the candidate abnormality probabilities in a field. This mean-probability aggregation was selected on validation data from maximum, top-*k* mean, noisy-OR and quality-gated alternatives, using the mean of the validation abnormality and high-grade average precisions, and the field operating thresholds were fixed on validation data. Field labels and scores are annotation-level quantities, not Bethesda slide reports.

### Statistical analysis

Performance was estimated separately for each task. Classification summaries included accuracy, macro-averaged F1 score, one-versus-rest AUROC and average precision; average precision weights precision by increments in recall, and positive prevalence is reported alongside it because precision depends on case mix. For the 95% CIs, we used bootstrapping with the strongest available grouping variable as the resampling unit: 500 resamples of source images for Cx22, 2,000 resamples of parent images for SIPaKMeD and 2,000 resamples of the 18 inferred groups for RIVA (seeds 2701 and 2702 for candidate AUROC and field average precision). Group resampling retained all observations within each sampled group. Ranges across the three CCCID models quantify model-fit variability and are not CIs. Bland–Altman limits of agreement^20^ were the mean model-minus-reference difference ± 1.96 s.d.

Four descriptive analyses were computed from the frozen predictions. For Cx22, we report the percentage of fields whose model count lay within *k* nuclei of the reference count. For SIPaKMeD and Herlev, we report the percentage of cells in which both compartments reached a Dice coefficient of at least 0.5 and at least 0.7. For Herlev, we computed the Spearman correlation between N:C and an ordinal squamous grade (normal superficial and intermediate, 0; mild dysplasia, 1; moderate, 2; severe, 3; carcinoma in situ, 4) separately for model and reference ratios, together with the AUROC of each ratio for separating the four abnormal classes from the three normal classes; these Herlev analyses are exploratory. For CCCID, we report the largest absolute difference between each broad-category probability and the sum of its fine-category probabilities, the percentage of stacks whose predicted fine category lay within the predicted broad category and the percentage of fine-category errors that remained within the correct broad category. No hypothesis tests or multiplicity-adjusted comparisons were performed.

### Software

Models were trained on Apple silicon with the Metal Performance Shaders backend of PyTorch. The analyses in this report were performed in Python (v.3.13.9) with NumPy (v.2.3.5), pandas (v.2.3.3), SciPy (v.1.18.0), scikit-learn (v.1.8.0) and PyTorch (v.2.12.1), and figures were produced with Matplotlib (v.3.10.8).

### Figure preparation

Microscopy examples are original dataset images and were not generated by an image model. Cx22 examples were selected at reference-count percentiles and CCCID examples by reference-class stack identifiers; SIPaKMeD examples were selected near the within-class median paired Dice coefficient and are illustrative. Their original 512-pixel source windows are shown at publication resolution; the recorded crop and bicubic reduction reproduce the archived 128-pixel model inputs exactly, and the evaluated mask contours are aligned to the same source-image coordinates. Only verified white preparation padding was omitted, and native aspect ratios were preserved. Quantitative plots were generated from the complete prediction tables. Fig. 1 is a schematic with values drawn from the reported results and introduces no data. Extended Data Fig. 1d reports an archived APACC candidate-validity control^14^.

### Use of AI-assisted tools

OpenAI Codex and Anthropic Claude were used to assist with manuscript drafting, cross-checking of code and reported results against the archived prediction tables, figure redesign and the reproducible preparation of figures and document files. No generative image model was used at any stage, and all microscopy pixels originate from the cited source datasets. No numerical result, CI or model output reported in this article was produced or altered by a language model. The authors reviewed and edited the content and take full responsibility for it.

## Data Availability

All source data are openly available from their original providers under their stated terms and must be obtained from them directly: Cx22 (https://github.com/LGQ330/Cx22), SIPaKMeD (https://www.cs.uoi.gr/~marina/sipakmed.html), Herlev (https://mde-lab.aegean.gr/index.php/downloads/), CCCID version 2 (https://doi.org/10.5281/zenodo.20807462), RIVA (https://zenodo.org/records/17288879) and APACC (https://osf.io/fp2xe). This preprint does not redistribute any source dataset. Split manifests and group identifiers, model configuration files and checkpoint hashes, complete per-unit prediction tables, model-selection and threshold-selection records, and the derived numeric tables behind every plotted panel are available from the corresponding authors on reasonable request.

https://www.cs.uoi.gr/~marina/sipakmed.html

https://mde-lab.aegean.gr/index.php/downloads/

https://doi.org/10.5281/zenodo.20807462

https://zenodo.org/records/17288879

https://osf.io/fp2xe

## Reporting summary

Further information on research design is available in the CLAIM 2024 checklist provided with this article.

## Data availability

All source data are openly available from their original providers under the terms shown and must be obtained from them directly; this preprint does not redistribute any source dataset.

- Cx22: https://github.com/LGQ330/Cx22 (non-commercial research and educational use, citation requested)
- SIPaKMeD: https://www.cs.uoi.gr/∼marina/sipakmed.html (experimental use with the request to cite)
- Herlev (DTU/Herlev Pap smear): https://mde-lab.aegean.gr/index.php/downloads/ (provider’s site terms)
- CCCID version 2: https://doi.org/10.5281/zenodo.20807462 (CC BY-NC 4.0)
- RIVA: https://zenodo.org/records/17288879 (CC BY-NC 4.0)
- APACC: https://osf.io/fp2xe (CC BY 4.0)

Split manifests and group identifiers for every reported partition, model configuration files and checkpoint hashes, complete per-unit prediction tables (including all errors and outliers), model-selection and threshold-selection records and the derived numeric tables behind every plotted panel permit recomputation of every value in Tables 2 and 3, Figs. 2–6 and Extended Data Figs. 1–5 from the original datasets. They are available from the corresponding authors on reasonable request. Microscopy panels in Figs. 2, 3 and 5 and Extended Data Fig. 4 reproduce Cx22^8^, SIPaKMeD^9^ and CCCID version 2^11^ images for non-commercial scholarly illustration with attribution; they remain under their providers’ terms and are not covered by the CC BY-NC-ND 4.0 license of this preprint. All other panels are plots of dataset-derived prediction tables or schematic drawings.

## Code availability

Model, analysis and figure-generation code are available from the corresponding authors on reasonable request.

## Acknowledgements

We thank the creators and maintainers of the Cx22, SIPaKMeD, Herlev, CCCID, RIVA and APACC datasets for making their data openly available.

## Author contributions

Korrine Lee and Rosa Jiwon Yoon contributed equally and are co-first authors. Korrine Lee: investigation, data curation, validation, visualization, writing – original draft, writing – review and editing. Rosa Jiwon Yoon: investigation, validation, formal analysis, writing – original draft, writing – review and editing. Dongyong Lee: conceptualization, methodology, software, formal analysis, data curation, visualization, funding acquisition, writing – review and editing. Robyn Sungmi Lee: conceptualization, supervision, project administration, writing – review and editing. All authors read and approved the final manuscript and agreed to its posting.

