## Supplementary File for "CytoLens: a modular deep learning system for quantitative cervical cytology"

**Extended Data Fig. 1 |** Nucleus detection and counting: agreement and error distribution

**Extended Data Fig. 2 |** Point-prompted segmentation on SIPaKMeD by class and parent image

**Extended Data Fig. 3 |** Transfer to Herlev: N:C agreement, input window and comparison with the earlier pipeline

**Extended Data Fig. 4 |** Hierarchical classification across focus and fine categories

**Extended Data Fig. 5 |** Abnormality scoring: aggregation, operating point and eight-category assignment

Reporting checklist: CLAIM 2024 (separate file, CytoLens_medRxiv_CLAIM2024_Checklist.pdf)

Extended Data figures share the design system, data sources and verification of the main figures. Source data for every plotted panel are available from the corresponding authors on reasonable request.

### Extended Data Fig. 1 | Nucleus detection and counting: agreement and error distribution

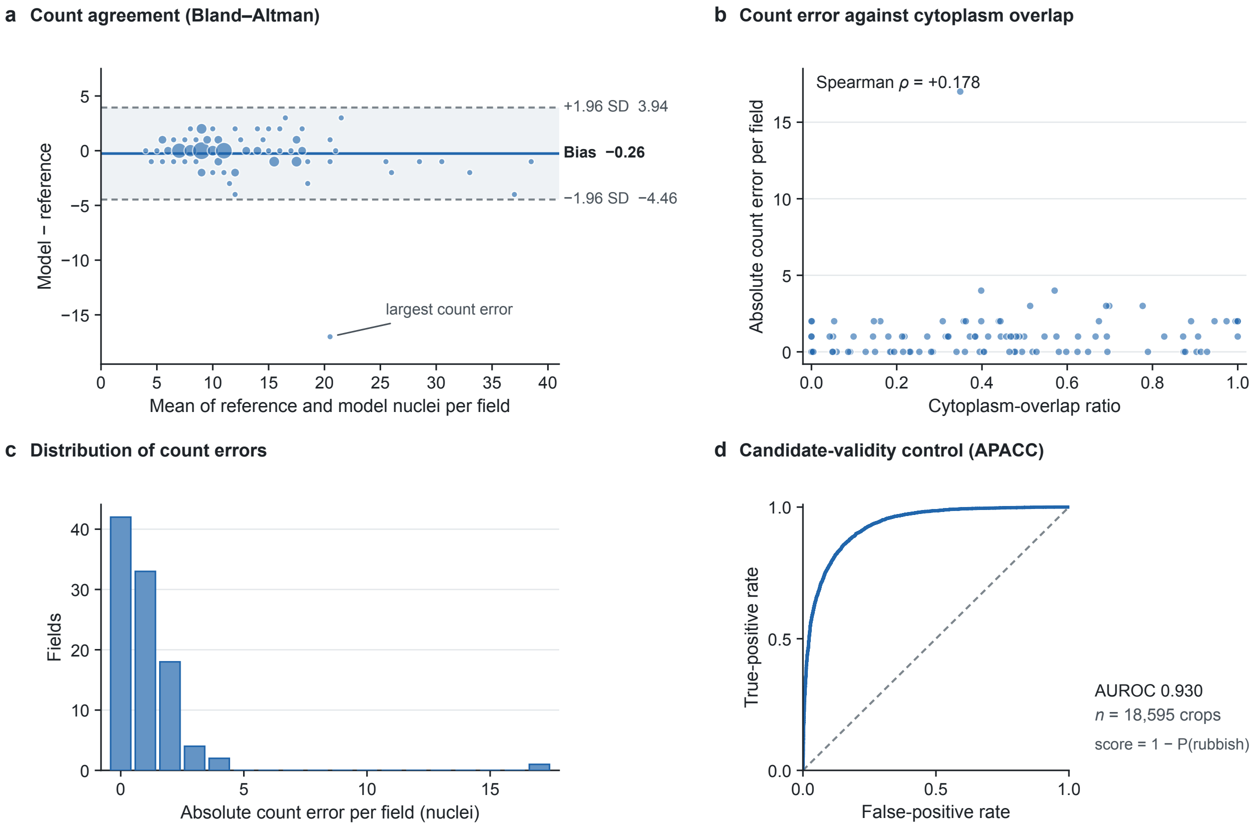


**a**, Bland–Altman plot of model minus reference nuclei per field against their mean (marker area proportional to the number of fields); bias, −0.26 nuclei per field; limits of agreement, −4.46 to 3.94 (bias ± 1.96 s.d.). The largest count error (17 nuclei) is labeled. **b**, Absolute count error per field against the cytoplasm-overlap ratio (Spearman $\rho$ = 0.178). **c**, Distribution of absolute count errors ($n$ = 100 fields). **d**, ROC curve of an archived APACC candidate-validity control that separates valid cell crops from debris with the score 1 − P(rubbish) ($n$ = 18,595 crops; AUROC, 0.930). APACC coarse labels do not establish cytologic abnormality. ROC, receiver operating characteristic; AUROC, area under the ROC curve.

### Extended Data Fig. 2 | Point-prompted segmentation on SIPaKMeD by class and parent image

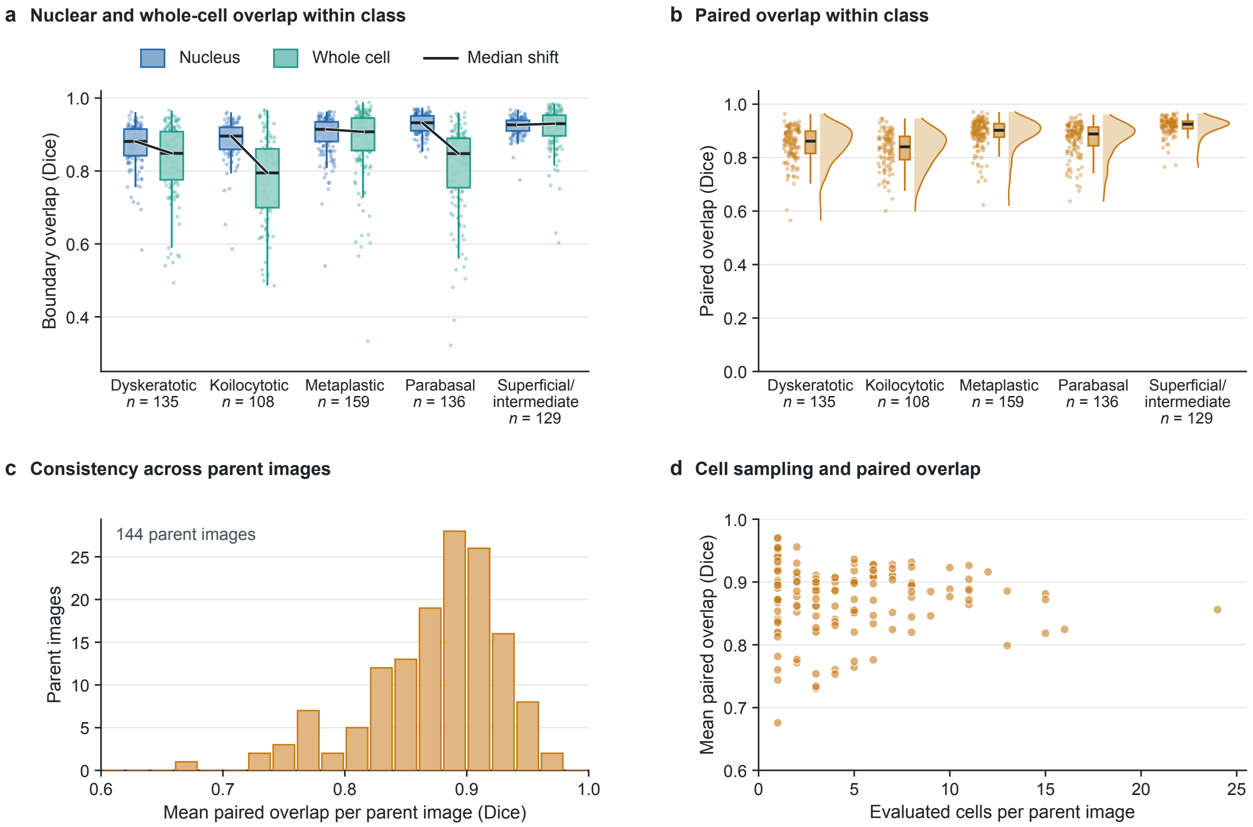


**a**, Nuclear (blue) and whole-cell (teal) Dice coefficients within each source class, with lines joining the two medians. Mean nuclear Dice coefficients were 0.871 (dyskeratotic), 0.884 (koilocytotic), 0.899 (metaplastic), 0.928 (parabasal) and 0.922 (superficial/intermediate). **b**, Paired Dice coefficient within each source class. **c**, Mean paired Dice coefficient per parent image ($n$ = 144). **d**, Number of evaluated cells per parent image versus the parent-image mean paired Dice coefficient. Boxes indicate quartiles and whiskers extend to the most extreme values within 1.5 × the interquartile range.

### Extended Data Fig. 3 | Transfer to Herlev: N:C agreement, input window and comparison with the earlier pipeline

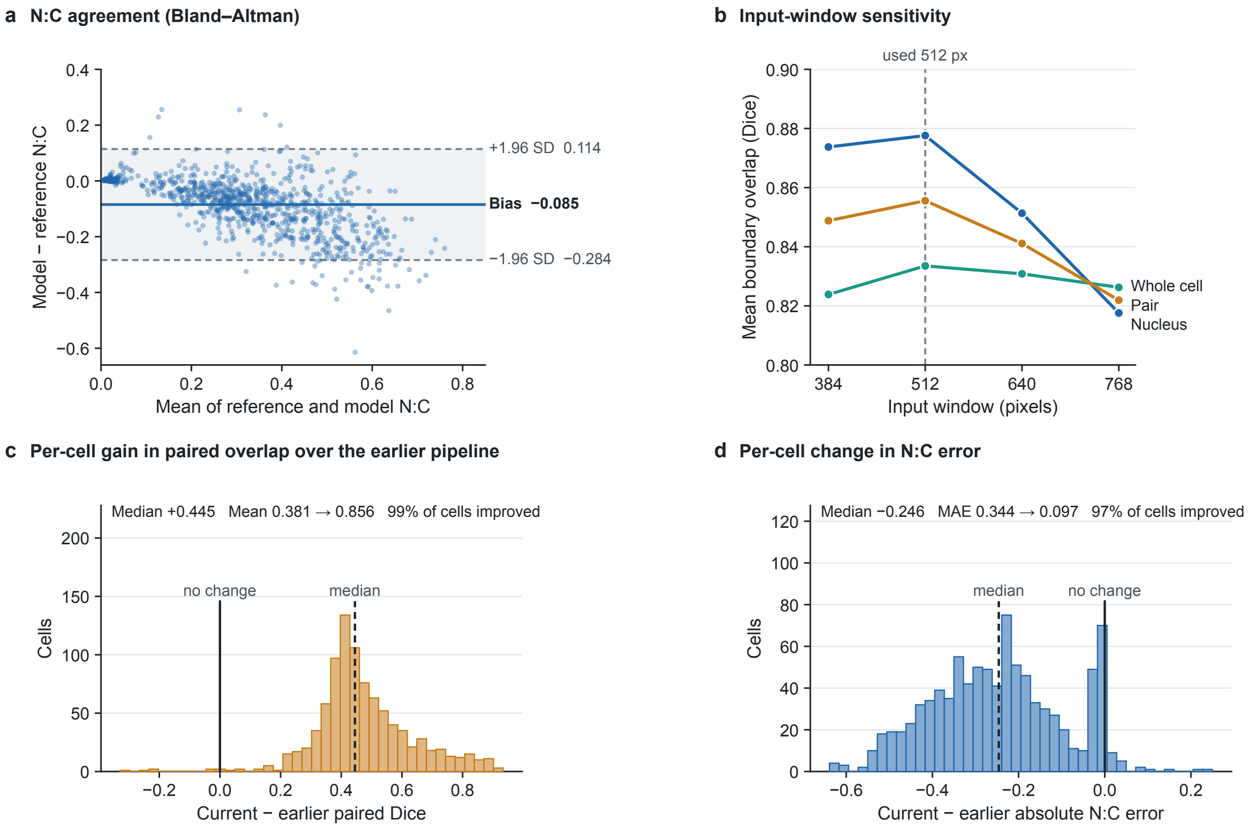


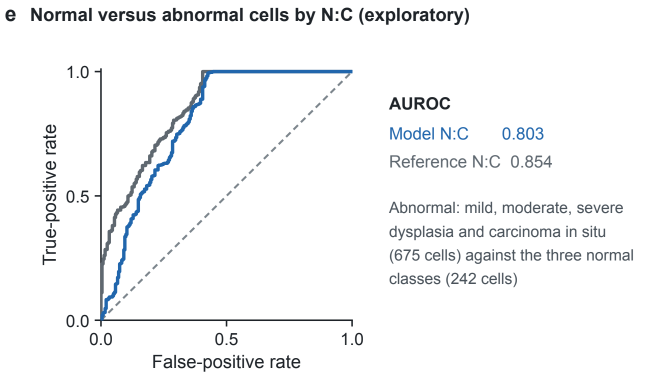


**f Median N:C by class**

| Class | Reference | Model | Cells |
|---|---|---|---|
| Normal columnar | 0.331 | 0.285 | 98 |
| Normal superficial | 0.010 | 0.014 | 74 |
| Normal intermediate | 0.029 | 0.031 | 70 |
| Mild dysplasia | 0.261 | 0.204 | 182 |
| Moderate dysplasia | 0.358 | 0.266 | 146 |
| Severe dysplasia | 0.467 | 0.351 | 197 |
| Carcinoma in situ | 0.611 | 0.437 | 150 |

**a**, Bland–Altman plot of model minus reference N:C ($n$ = 917 cells); mean difference, −0.085; limits of agreement, −0.284 to 0.114. **b**, Mean Dice coefficients for input windows of 384–768 pixels; the 512-pixel window was fixed on SIPaKMeD validation data before Herlev was analyzed. **c,d**, Per-cell change from the earlier Cx22-trained CytoLens segmentation pipeline to the current SIPaKMeD-trained pipeline. The mean paired Dice coefficient increased from 0.381 to 0.856 and improved in 99% of cells (**c**); the N:C mean absolute error decreased from 0.344 to 0.097 and improved in 97% of cells (**d**). Training source, input window and thresholds changed together. **e**, Exploratory ROC curves for separating abnormal cells (mild, moderate and severe dysplasia and carcinoma in situ; $n$ = 675) from normal cells ($n$ = 242) with N:C alone; AUROC, 0.803 (model) and 0.854 (reference). **f**, Median reference and model N:C by class. ROC, receiver operating characteristic; AUROC, area under the ROC curve.

### Extended Data Fig. 4 | Hierarchical classification across focus and fine categories

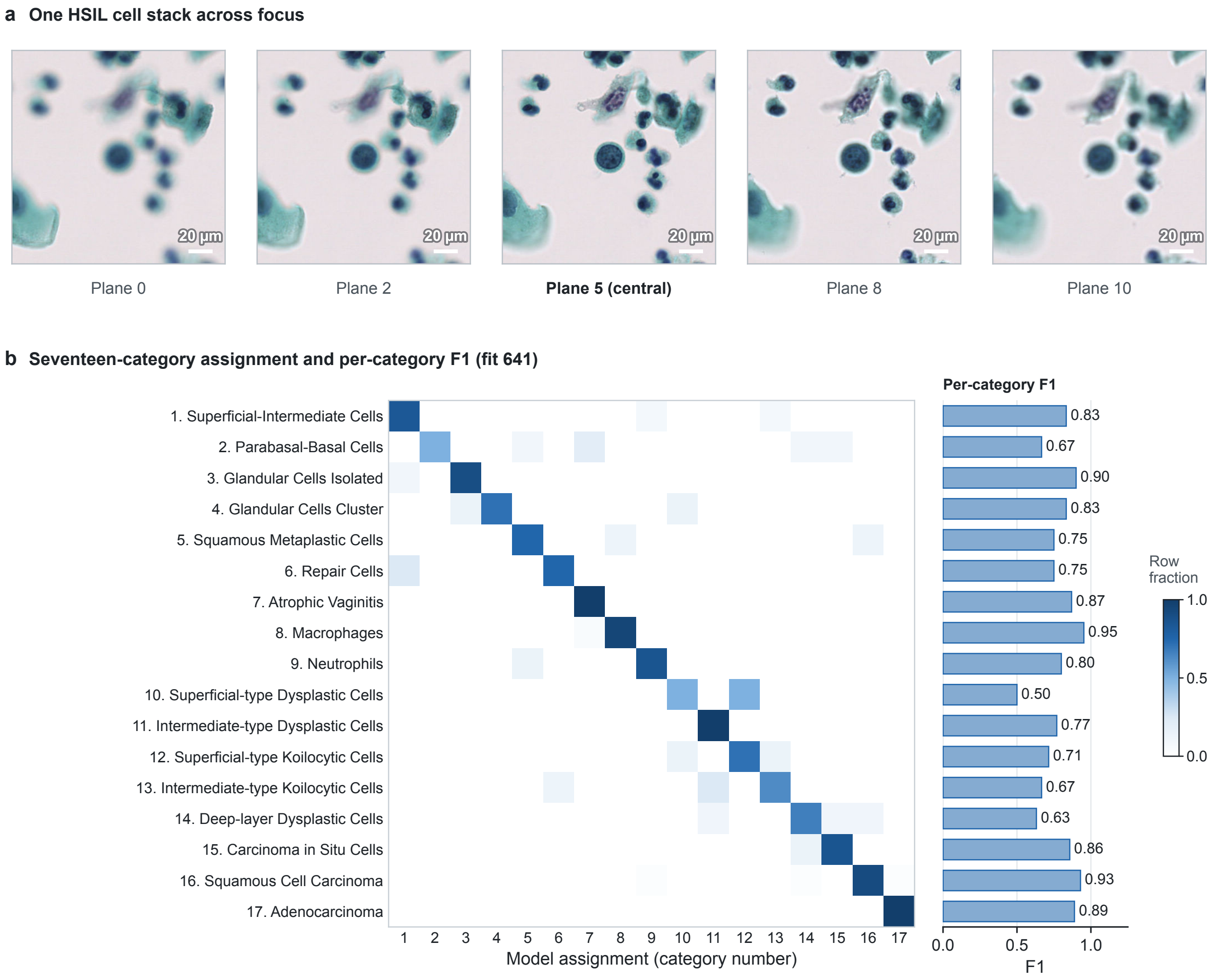


**a**, Five of the 11 focal planes of one HSIL test stack (planes 0, 2, 5, 8 and 10; scale bars, 20 µm); the model averages the logits of each output head over all 11 planes. **b**, Seventeen-class confusion matrix (row fractions) and per-class F1 scores for the primary model ($n$ = 176 stacks). Classes 1–9 belong to NILM, 10–13 to LSIL, 14–15 to HSIL and 16–17 to SCC/other. Accuracy and macro-averaged F1 score were 0.830 and 0.783 for this model and 0.850 and 0.820 on average across the three models.

**Extended Data Fig. 5 | Abnormality scoring: aggregation, operating point and eight-category assignment**

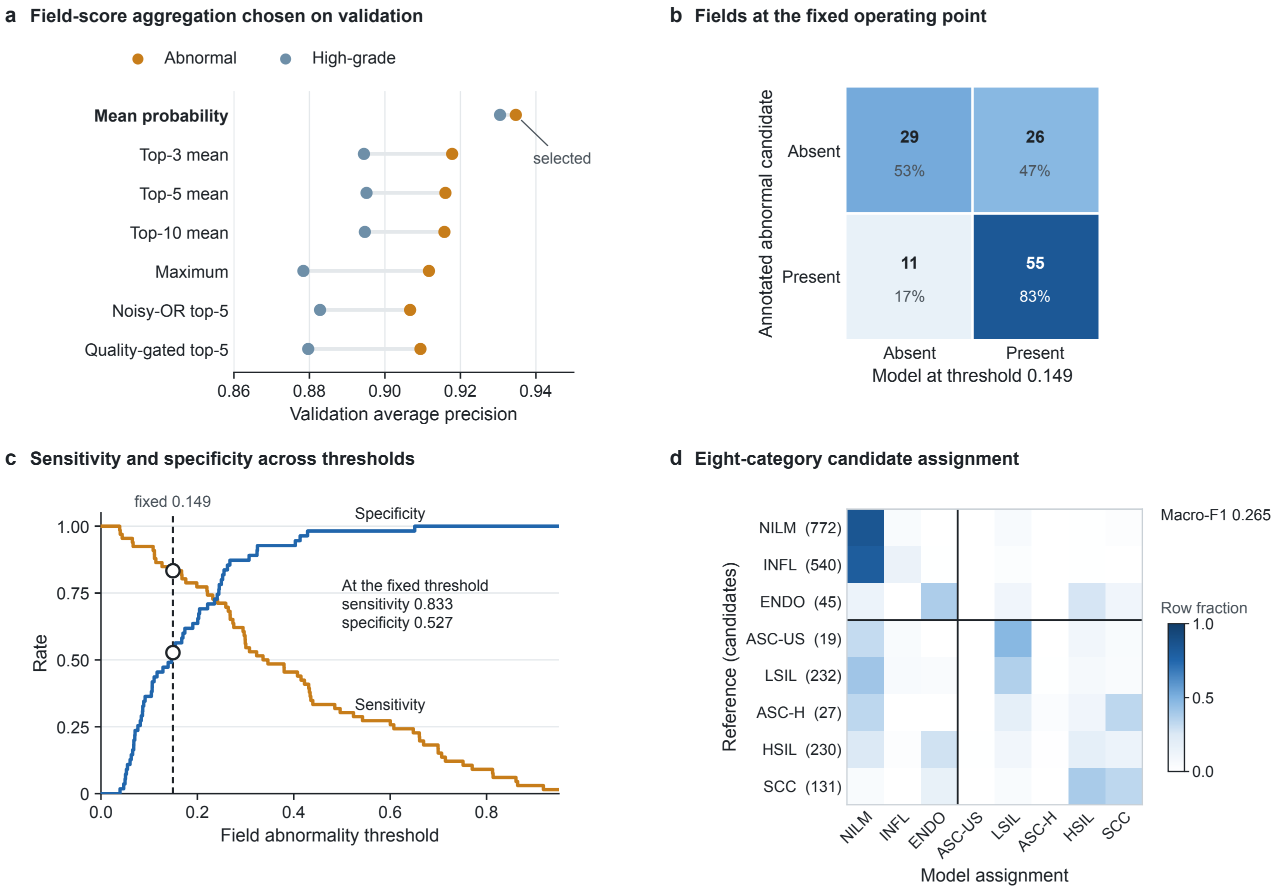


**a**, Validation average precision of candidate-to-field aggregation rules for the abnormal (amber) and high-grade (slate) endpoints; the mean-probability rule was selected on validation data before testing. **b**, Test fields at the fixed abnormality threshold of 0.149 (55 true positives, 26 false positives, 11 false negatives and 29 true negatives; sensitivity, 0.833; specificity, 0.527). **c**, Sensitivity and specificity across field thresholds, with the fixed operating point marked; the high-grade operating point had a sensitivity of 0.692 and a specificity of 0.805 (Table 3). **d**, Eight-category assignment of candidates (row fractions; candidate counts in parentheses); macro-averaged F1 score, 0.265. Lines separate non-lesion labels from lesion grades.
